# Predictors of survival in patients with ischemic stroke and active cancer: A prospective, multicenter, observational study

**DOI:** 10.1101/2023.05.08.23289699

**Authors:** Yasufumi Gon, Manabu Sakaguchi, Hiroshi Yamagami, Soichiro Abe, Hiroyuki Hashimoto, Nobuyuki Ohara, Daisuke Takahashi, Yuko Abe, Tsutomu Takahashi, Takaya Kitano, Shuhei Okazaki, Kenichi Todo, Tsutomu Sasaki, Satoshi Hattori, Hideki Mochizuki, SCAN Study Investigators

**Affiliations:** Department of Neurology, Osaka University Graduate School of Medicine, Osaka, Japan; Department of Neurology, Osaka General Medical Center, Osaka, Japan; Department of Neurology, National Hospital Organization Osaka National Hospital, Osaka, Japan; Department of Neurology, National Cerebral and Cardiovascular Center, Osaka, Japan; Department of Neurology, Osaka Rosai Hospital, Osaka, Japan; Department of Neurology, Kobe City Medical Center General Hospital, Hyogo, Japan; Department of Neurology, National Hospital Organization Osaka Minami Medical Center, Osaka, Japan; Department of Neurology, Yodogawa Christian Hospital, Osaka, Japan; Department of Neurology, Hoshigaoka Medical Center, Osaka, Japan; Department of Integrated Medicine, Biomedical Statistics, Osaka University of Graduate School of Medicine, Osaka, Japan

**Keywords:** ischemic stroke, active cancer, D-dimer, cryptogenic stroke, distant metastasis

## Abstract

**Background:** Patients with ischemic stroke and active cancer have a poor prognosis; however, supporting evidence remains limited.

**Methods:** We conducted a prospective, multicenter, observational study in Japan including patients with acute ischemic stroke and active cancer to investigate the prognostic factors. We followed up the patients for 1 year after stroke onset. The patients were divided into two groups according to cryptogenic stroke and known etiologies (small vessel occlusion, large artery atherosclerosis, cardioembolism, other determined etiology) and survival was compared. The hazard ratios (HRs) and 95% confidence intervals (CIs) for mortality were calculated using Cox regression models.

**Results:** We identified 135 eligible patients (39% women; median age, 75 years). Of these, 51% had distant metastasis. A total of 65 (48%) and 70 (52%) patients had cryptogenic stroke and known etiologies, respectively. Patients with cryptogenic stroke had significantly shorter survival than those with known etiologies (HR [95% CI], 3.11 [1.82–5.25]). The multivariate Cox regression analysis revealed that distant metastasis, plasma D-dimer levels, deep venous thrombosis and/or pulmonary embolism complications at stroke onset were independent predictors of mortality after adjusting for potential confounders. Cryptogenic stroke was associated with prognosis in univariate analysis but not significant in multivariate analysis. The plasma D-dimer levels stratified the prognosis of patients with ischemic stroke and active cancer.

**Conclusions:** The prognosis of patients with acute ischemic stroke and active cancer varies considerably depending on stroke mechanism, distant metastasis, and coagulation abnormalities. Coagulation abnormalities are crucial in determining the prognosis of such patients.

**What Is New?:**

- We conducted a prospective, multicenter, observational study in Japan to determine the prognostic survival factors in patients with acute ischemic stroke and active cancer.
- Distant metastasis, plasma D-dimer levels, deep venous thrombosis and/or pulmonary embolism were independent predictors of mortality after adjusting for potential confounders.
- Patients with known stroke etiologies and mild coagulation abnormalities had a favorable prognosis, whereas those with cryptogenic stroke and severe coagulation abnormalities had a poor outcome.

**What Are the Clinical Implications?:**

- The prognosis of patients with acute ischemic stroke and active cancer varies considerably depending on stroke mechanism, distant metastasis, and coagulation abnormalities.
- Patients with known stroke etiologies and mild coagulation abnormalities often have a favorable prognosis; therefore, we should not stop stroke therapy because of active cancer.
- Patients with cryptogenic stroke and severe coagulation abnormalities often have poor outcomes; consequently, we should thoroughly discuss with the oncologist to determine a treatment plan.

## Introduction

Patients with cancer are at increased risk of cerebrovascular diseases [1,2]. Stroke is a significant concern for patients with cancer since it can interrupt cancer treatments and worsen survival outcomes [1–5]. Management of cancer and stroke is an important aspect of clinical practice.

Patients with ischemic stroke and active cancer experience poor survival outcomes. Lee et al. investigated 268 patients with ischemic stroke and active cancer using data from the OASIS-Cancer study and reported a median survival of 109 days [6]. Navi et al. analyzed 263 patients with ischemic stroke and active cancer and found a median survival of 84 days for 230 patients with complete follow-up [7]. Shin et al. investigated 93 patients with cryptogenic stroke and active cancer and showed a median survival of 62 days [8]. Other studies compared ischemic stroke outcomes in patients with and without active cancer, suggesting that ischemic stroke patients with active cancer have poorer outcomes than those without [9–14]. Overall, patients with ischemic stroke and active cancer are known to have poor survival outcomes. However, most of these studies are retrospective [7–12,14], and limited data are available concerning prospective studies [6,13].

The prognostic indicators of survival are poorly understood in patients with ischemic stroke and active cancer [15]. Previous studies have suggested that distant metastasis, diabetes mellitus, plasma D-dimer levels, cryptogenic stroke, and pre-stroke modified Rankin Scale (mRS) were associated with mortality [8,16–18]. Among these factors, cryptogenic stroke is a common ischemic stroke subtype in patients with active cancer [16,19–21]. It is also known that plasma D-dimer levels are high in cancer patients with cryptogenic stroke [19,20]. Thus, the fact that both cryptogenic stroke and plasma D-dimer levels predict prognosis may reflect underlying coagulation abnormalities. However, the relationship between cryptogenic stroke and plasma D-dimer levels in predicting cancer-related stroke prognosis has been underexplored.

Based on this background, we conducted a prospective, multicenter, observational study of patients with ischemic stroke and with cancer (ischemic Stroke in patients with CAncer and Neoplasia [SCAN]) in Japan. The SCAN study was designed to assess the clinical characteristics of cancer-related strokes. In the present study, we analyzed the survival of patients with acute ischemic stroke and active cancer according to the stroke subtype, using data from the SCAN study.

## Methods

### Data Availability

The data that support the findings of this study are available from the corresponding author upon reasonable request.

### Standard Protocol Approvals, Registrations, and Patient Consents

This study complied with the Declaration of Helsinki for investigations involving humans and was approved by the Institutional Review Board of Osaka University Hospital (approval number: 15346-10). Written informed consent was obtained from all study patients.

### Study Design and Population

The SCAN study is a prospective, multicenter, observational study conducted at nine hospitals in Japan, between June 2016, and December 2021 (Osaka University Hospital, Osaka General Medical Center, National Cerebral and Cardiovascular Center Hospital, National Hospital Organization Osaka National Hospital, Osaka Rosai Hospital, Kobe City Medical Center General Hospital, National Hospital Organization Osaka Minami Medical Center, Yodogawa Christian Hospital, and Hoshigaoka Medical Center). All hospitals included the SCAN study function as regional stroke centers. The inclusion criteria were as follows: (1) acute ischemic stroke within 14 days following symptom onset, (2) age ≥20 years, and (3) active cancer at stroke onset. Active cancer was defined as a diagnosis of cancer, either treated in the last six months before admission or untreated, or metastatic disease. The SCAN study aimed to enroll approximately 250 patients over a 5-year study period, with a 1-year follow-up from the enrollment date to investigate the prognosis of patients with acute ischemic stroke and active cancer.

### Clinical Variables

The following baseline variables were obtained from the SCAN database: age, sex, hypertension, dyslipidemia, diabetes mellitus, smoking, atrial fibrillation, past stroke, time from symptom onset to admission, antithrombotic (antiplatelet and/or anticoagulant) medication before and after stroke onset, stroke subtype, National Institute of Health Stroke Scale (NIHSS), pre-stroke mRS, deep venous thrombosis and/or pulmonary embolism (DVT/PE) complications at stroke onset, infarct pattern of brain imaging, use of recombinant-tissue plasminogen activator (rt-PA), mechanical thrombectomy, and cancer-related data including cancer type, distant metastasis, adenocarcinoma, and cancer treatment (cancer surgery, chemotherapy, and radiotherapy). We also collected the recurrent stroke and major bleeding information during the observation period. Stroke recurrence was defined as a new neurological deficit together with corresponding evidence of acute ischemia or hematoma on brain imaging (computed tomography and/or magnetic resonance imaging). Major bleeding was defined as clinically overt bleeding that was accompanied by one or more of the following: a decrease in hemoglobin levels of at least 2 g/dL or the requirement of a transfusion with at least 2 units of packed red blood cells; symptomatic bleeding requiring immediate treatment that occurred at a critical site, such as intracranial, intraocular, intraspinal, intra-articular, intramuscular with compartment syndrome, pericardial or retroperitoneal; or bleeding that was fatal. Blood samples were collected immediately after admission before any treatment started, and plasma D-dimer and high sensitivity C-reactive protein (hsCRP) levels were quantified.

Subtypes of ischemic stroke were classified according to the Trial of Org 10172 in Acute Stroke Treatment (TOAST) criteria: small vessel occlusion, large artery atherosclerosis, cardioembolism, other determined etiology, and stroke of undetermined etiology [24]. All the patients underwent blood tests, 24-hour electrocardiographic monitoring at least 7 days after admission, carotid ultrasonography, and brain imaging (computed tomography and/or magnetic resonance imaging). Transthoracic and transesophageal echocardiography was performed as necessary. We determined that patients had a cryptogenic stroke if the cause could not be determined with any degree of confidence [24]. Patients with no apparent cause of ischemic stroke other than malignancies were categorized as having a cryptogenic stroke.

### Follow Up Duration and Outcomes

The observation period was one year after the diagnosis of ischemic stroke. All the patients were followed-up via telephone or outpatient visits. The incidence of death, stroke recurrence, and major bleeding were investigated.

### Statistical Analyses

Descriptive statistics were presented using the median and interquartile range (IQR) for continuous variables and as proportions and counts for categorical variables.

First, we summarized the baseline characteristics of the cohort. Next, we divided the patients into two groups based on the cryptogenic stroke and known etiologies (small vessel occlusion, large artery atherosclerosis, cardioembolism, and other determined etiology) and verified the survival rates against previously reported findings [7,8]. Survival rates for all-cause mortality were calculated using the Kaplan-Meier method. Finally, we estimated hazard ratios (HRs) and 95% confidence intervals (CIs) for mortality using univariate and multivariate Cox proportional hazard models. Based on clinical experience, we selected the following variables as potential confounders: age, sex, hypertension, dyslipidemia, diabetes mellitus, atrial fibrillation, pre-mRS ≤2, NIHSS score, distant metastasis, cryptogenic stroke, recurrent stroke, major bleeding, past stroke, pre-stroke antithrombotic use, pre-stroke cancer treatment status, DVT/PE complications at stroke onset, and plasma D-dimer levels. In the multivariate analysis, we developed two models to identify independent predictors of prognosis: the forced entry method for all potential confounders (Model 1) and the stepwise selection method to address the possibility of multicollinearity (Model 2). Since we were interested in cryptogenic stroke as a prognostic marker for survival, the stroke mechanism was forced to be included in Model 2, and other factors were selected using a stepwise procedure with Akaike’s information criterion (AIC) [25]. Recurrent stroke and major bleeding were treated as time-varying variables. Plasma D-dimer levels were divided into quartiles, and the first quartile was used as a reference.

The cumulative incidence of recurrent stroke and major bleeding during the observation period was calculated using the Fine and Gray model [26], with death as a competing risk. The incidence was compared with the use of antithrombotic medications.

Statistical analysis was performed using the R software (https://cran.r-project.org/). The level of significance was set at *P* < 0.05. All tests were two tailed.

## Results

### Study Population and Characteristics of the Patients according to Stroke Subtypes

A total of 135 patients were included in the analysis. **Table 1** lists the cancer types of the included patients and whether distant metastasis were present. The most common types of cancers were lung (16%), colorectal (14%), pancreatic (13%), prostate (9%), and gastric (8%) cancers. Distant metastasis was observed in 51% (n = 69) of the patients.

**Table 1.**
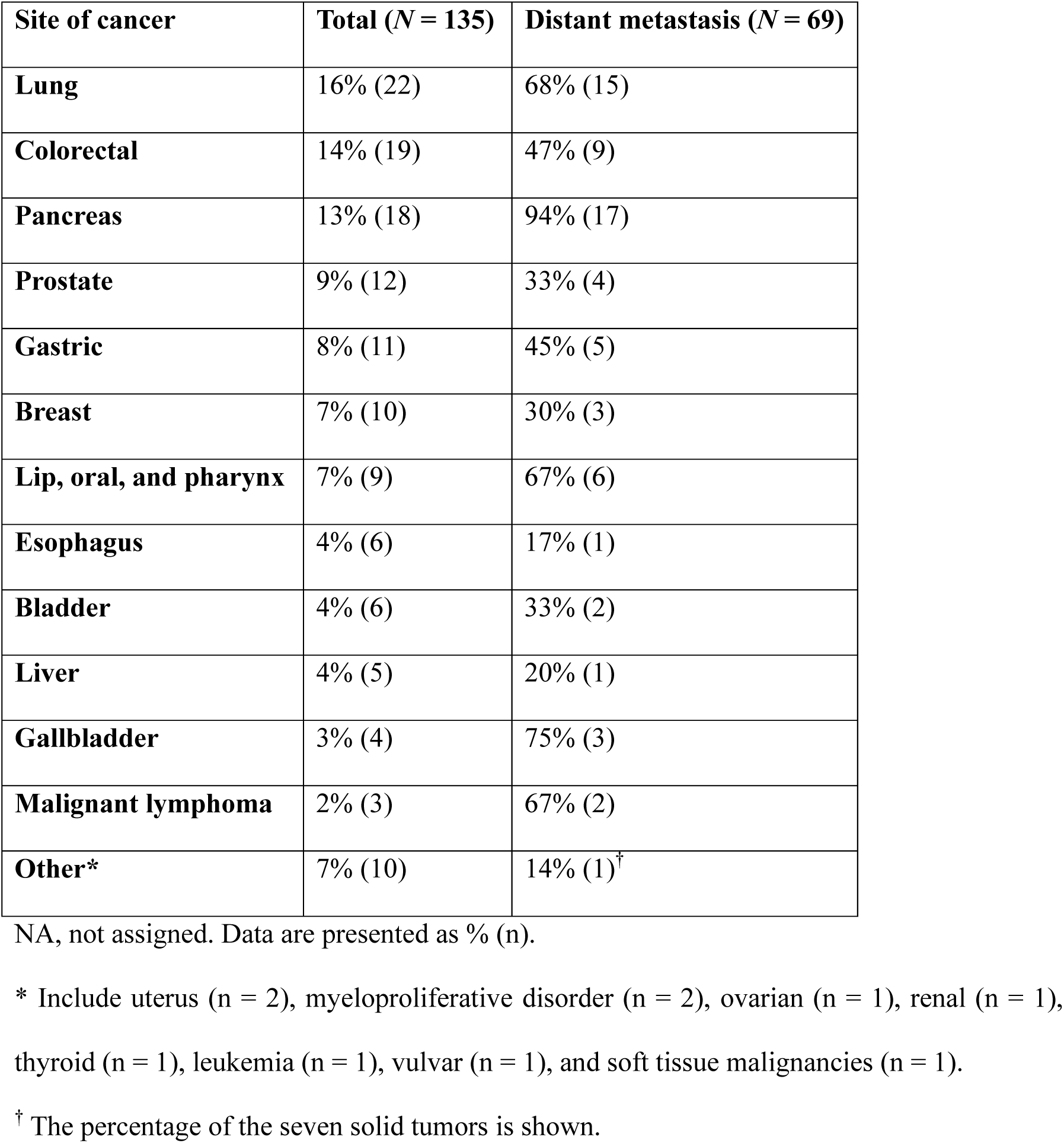
Index of cancer

**Table 2** summarizes the characteristics of the study cohort. Overall, the median (IQR) age was 75 (69–81) years, and 39% of the patients were women. The median (IQR) time from symptom onset to admission was 0 (0–2) days. Antithrombotic medications were used in 30% of the patients before stroke and 82% after ischemic stroke (breakdown of antithrombotic medications after stroke is shown in **Table S1**). Cryptogenic stroke was the most common stroke subtype (48%), and other determined etiology was the least common (7%). The median NIHSS (IQR) score was 4 (2–10). Ten patients (7%) had DVT/PE at stroke onset. Thrombolysis by rt-PA and mechanical thrombectomy were performed in 7% and 10% of the patients, respectively. The frequency of the patients undergoing cancer treatment was 76%, and approximately half of them received chemotherapy. The median (IQR) levels of plasma D-dimer and hsCRP were 3.65 (1.41–11.72) μg/mL and 1.06 (0.18–3.95) mg/dL, respectively.

**Table 2.**
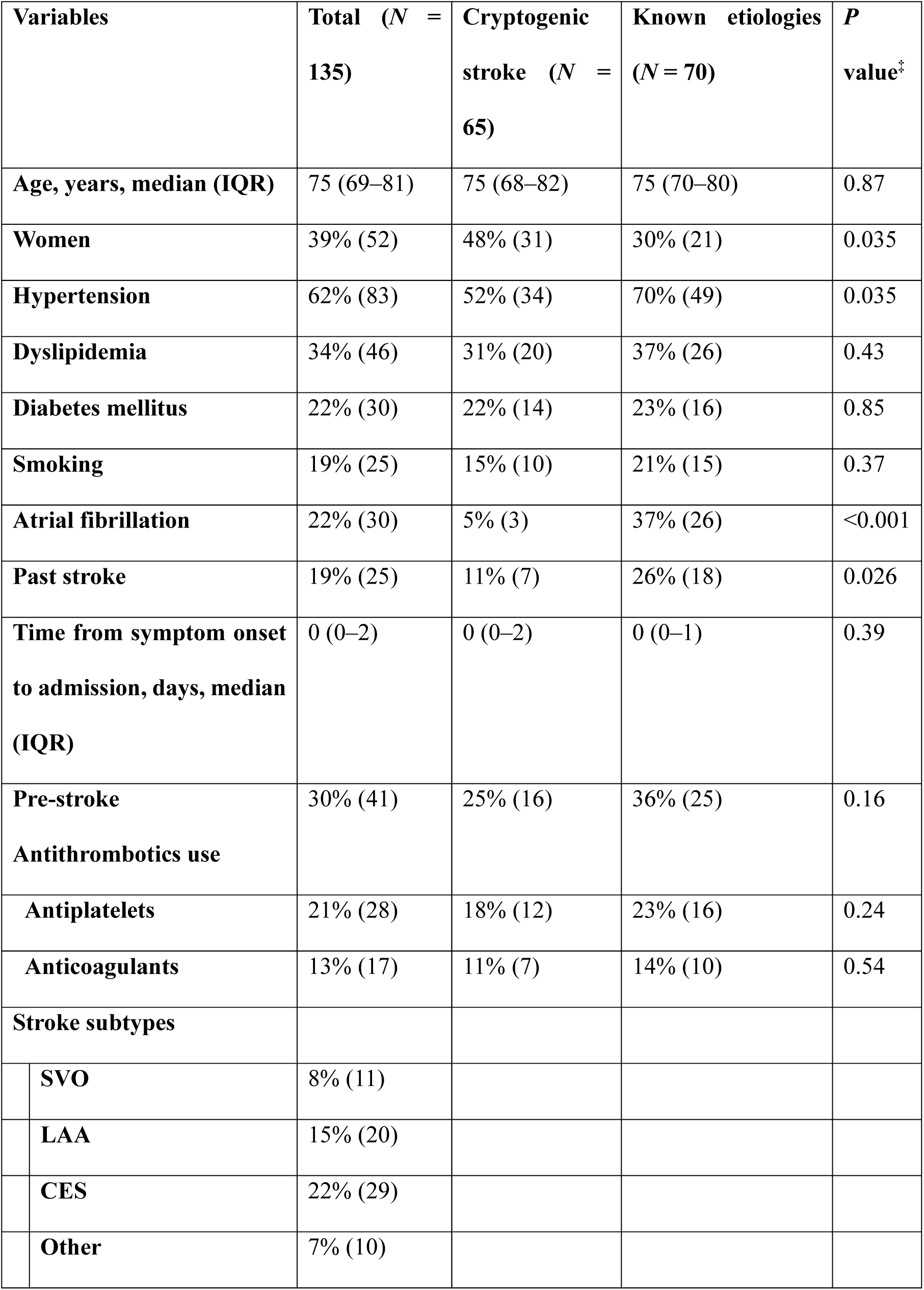
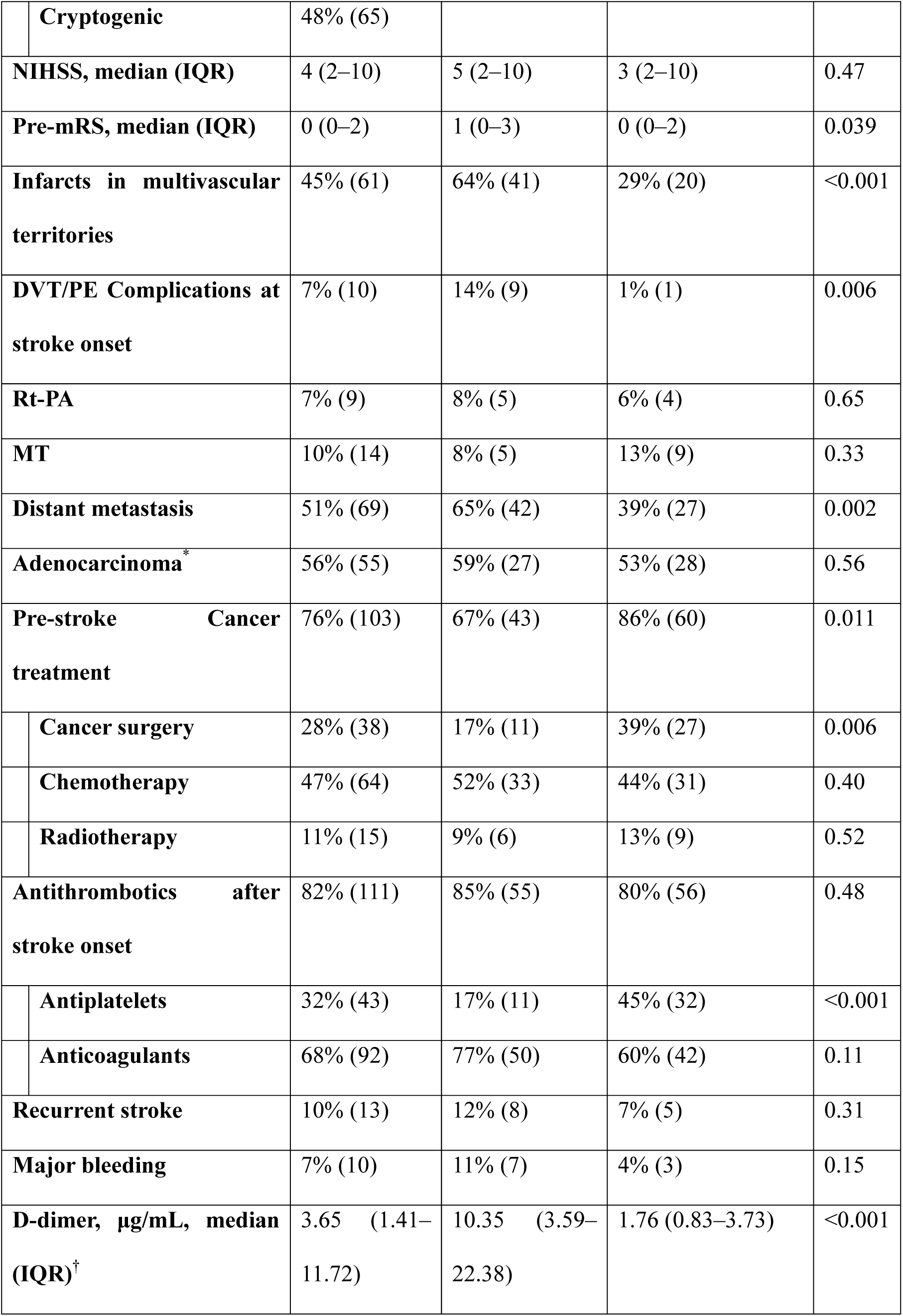
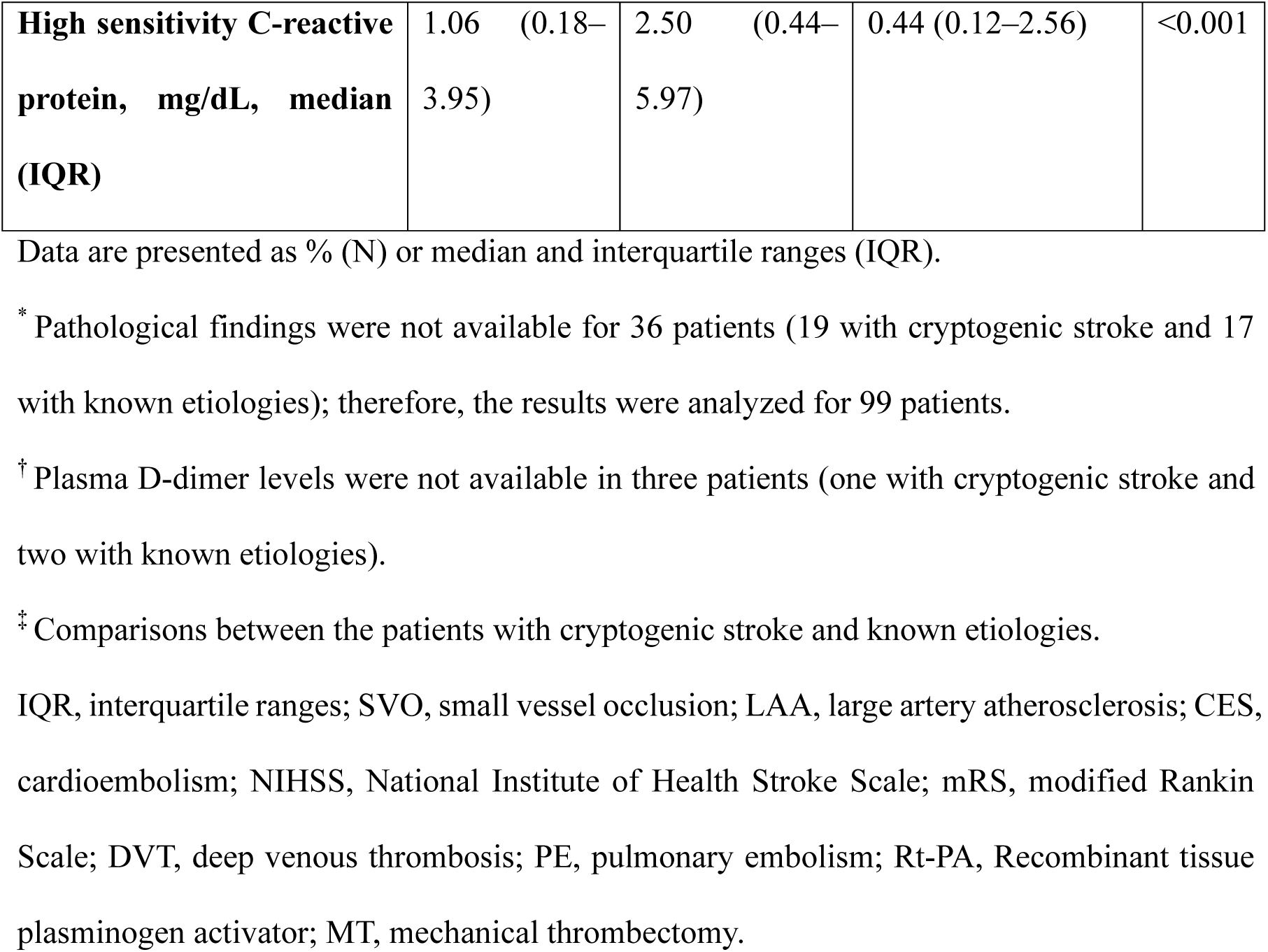
Characteristics of the patients according to stroke subtypes

Compared to the patients with known etiologies, those with cryptogenic stroke were more frequent in women, infarcts in multiple vascular territories, DVT/PE complications at stroke onset, and distant metastasis, while less frequent in those with hypertension, atrial fibrillation, past stroke, cancer treatment, and antiplatelet medication after stroke. Pre-stroke functional status was worse in patients with cryptogenic stroke compared with those with known etiologies (pre-mRS [IQR], 1 [0–3] vs. 0 [0–2], *P* = 0.039). The cryptogenic stoke group had higher plasma D-dimer levels (10.35 [3.59–22.38] μg/mL vs. 1.76 [0.83–3.73] μg/mL, *P* < 0.001) and serum hsCRP levels (2.50 [0.44–5.97] mg/dL vs. 0.44 [0.12–2.56] mg/dL, *P* < 0.001) compared to the known etiologies group.

### Survival Outcomes and Prognostic Factors

Among the 135 patients enrolled, 133 had completed 1-year follow-up: two cases were censored at the end of the follow-up. The median (IQR) survival time for all patients was 365 (71–365) days. The survival rates (95% CI) at 3 and 12 months were 70 (62–78) % and 54 (46– 63) %, respectively. **Figure 1** shows the difference in survival rates between patients with cryptogenic stroke and known etiologies. Patients with cryptogenic stroke had shorter survival times than those with known etiologies (113 [43–365] days vs. 365 [204–365] days, *P* < 0.001; HR [95% CI], 3.11 [1.82–5.25]).

**Figure 1.**
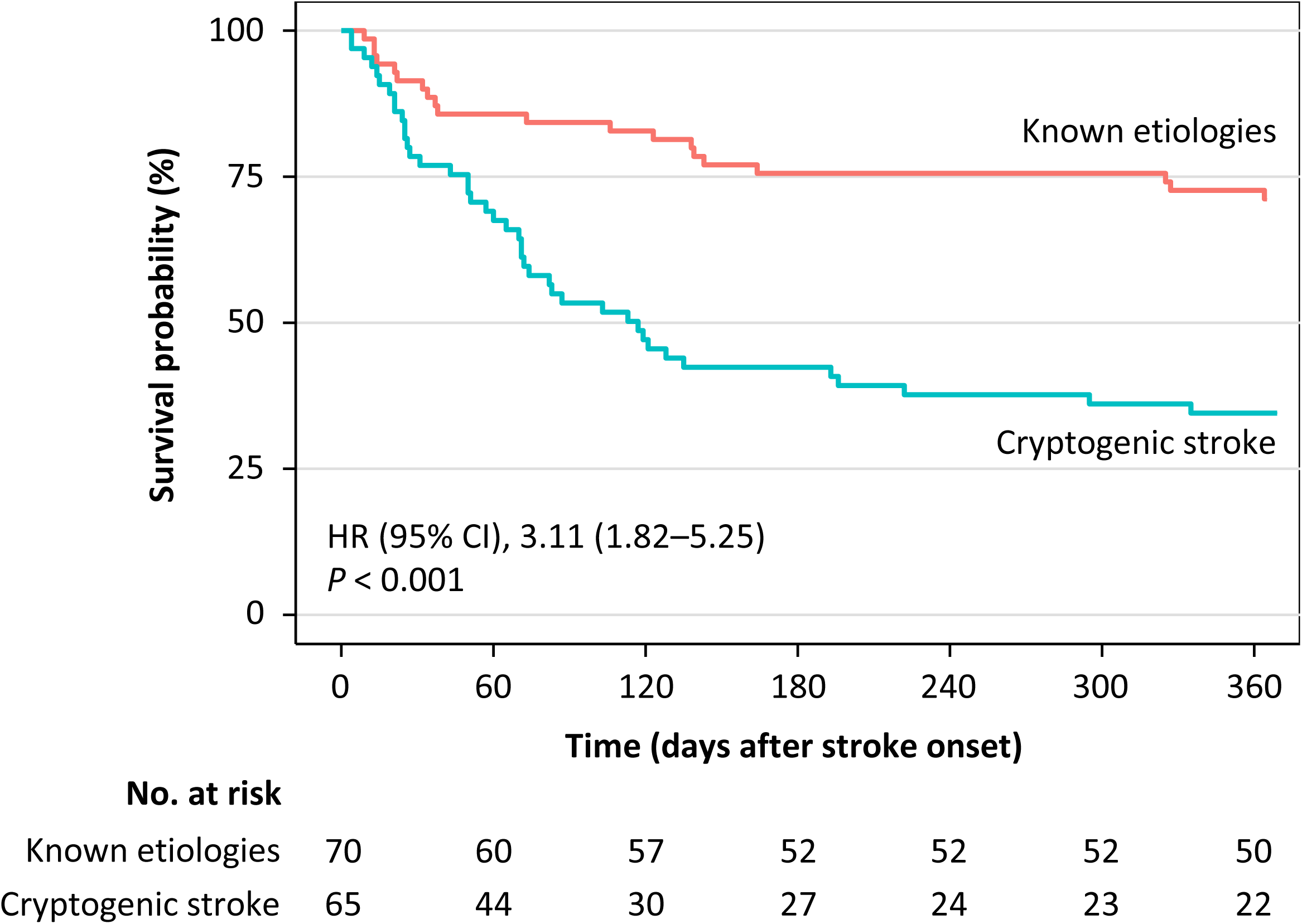
Kaplan-Meier survival curves by stroke mechanism

**Table 3** summarizes the Cox regression analyses. Univariate analysis revealed that hypertension, atrial fibrillation, cryptogenic stroke, NIHSS score, DVT/PE complications at stroke onset, distant metastasis, pre-stroke cancer treatment, recurrent stroke, and plasma D-dimer levels were associated with survival outcomes. Multivariate analysis revealed that DVT/PE complications at stroke onset and distant metastasis were significantly associated with mortality in Model 1, the forced entry method for all potential confounders. In Model 2, which selected variables using a stepwise procedure with AIC, DVT/PE complications at stroke onset, distant metastasis, and plasma D-dimer levels were independent predictors of prognosis. Cryptogenic stroke was associated with outcomes in univariate analysis but not significant in multivariate analysis. These results suggest a strong relationship between cancer progression, the degree of cancer-associated coagulopathy, and prognosis in patients with ischemic stroke and active cancer.

**Table 3.**
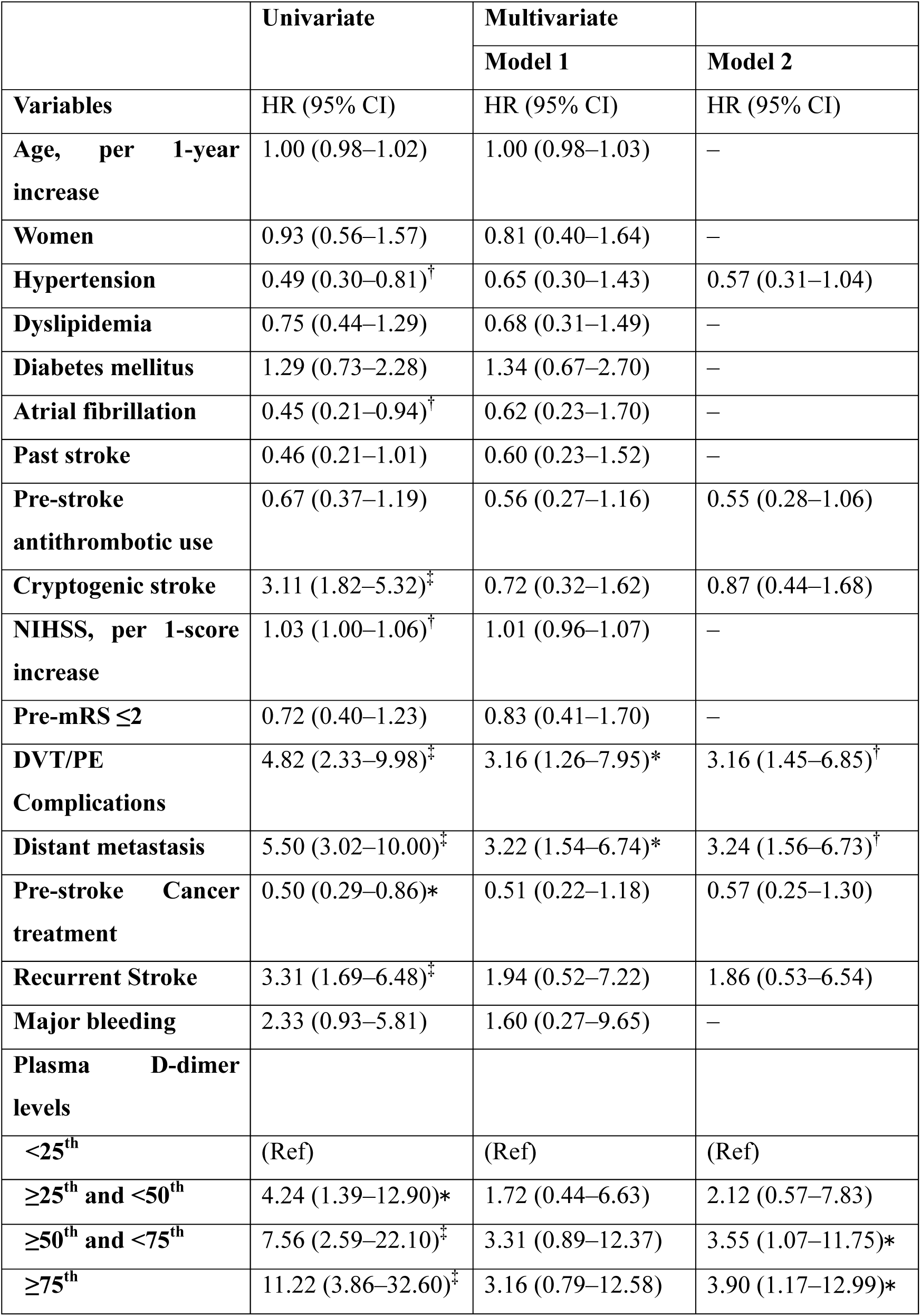
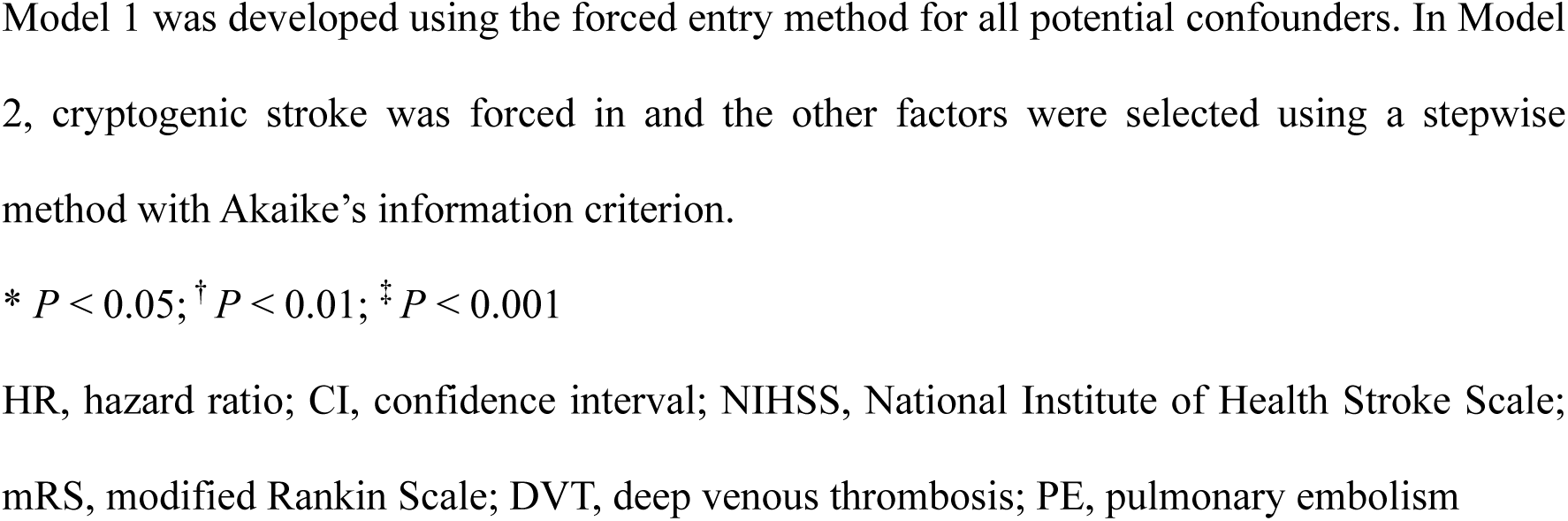
Hazard ratios for mortality using Cox regression model (univariate and multivariate)

### Recurrent Stroke and Major Bleeding during the Observation Period

The cumulative incidence (95% CI) of recurrent stroke and major bleeding were 10 (5–15) % and 7 (3–12) %, respectively. **Figure 2** shows the cumulative incidence of recurrent stroke between the groups treated with and without antithrombotic medications after stroke. The recurrent stroke was significantly lower in the group with antithrombotic agents than in those without (7 [3–13] % vs. 21 [8–39] %, *P* = 0.04; HR [95% CI], 0.32 [0.11–0.97]). **Figure 3** shows the cumulative incidence of major bleeding between the groups. There was no significant difference in major bleeding between the groups treated with and without antithrombotic medications (8 [1–23] % vs. 6 [3–12] %, *P* = 0.86; HR [95% CI], 0.87 [0.18–4.10]).

**Figure 2.**
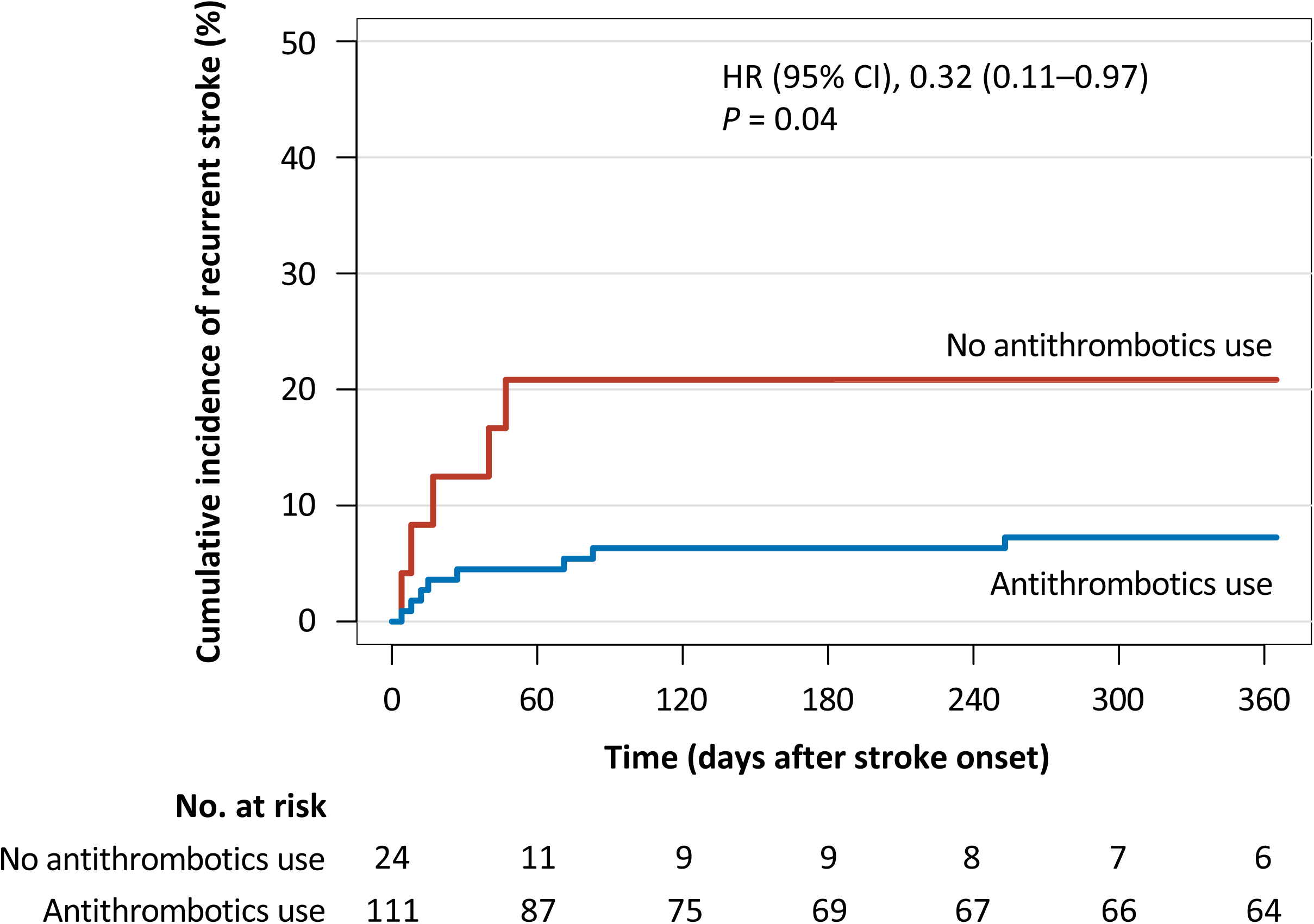
Cumulative incidence of recurrent stroke by antithrombotic agents use HR, hazard ratio; CI, confidence interval

**Figure 3.**
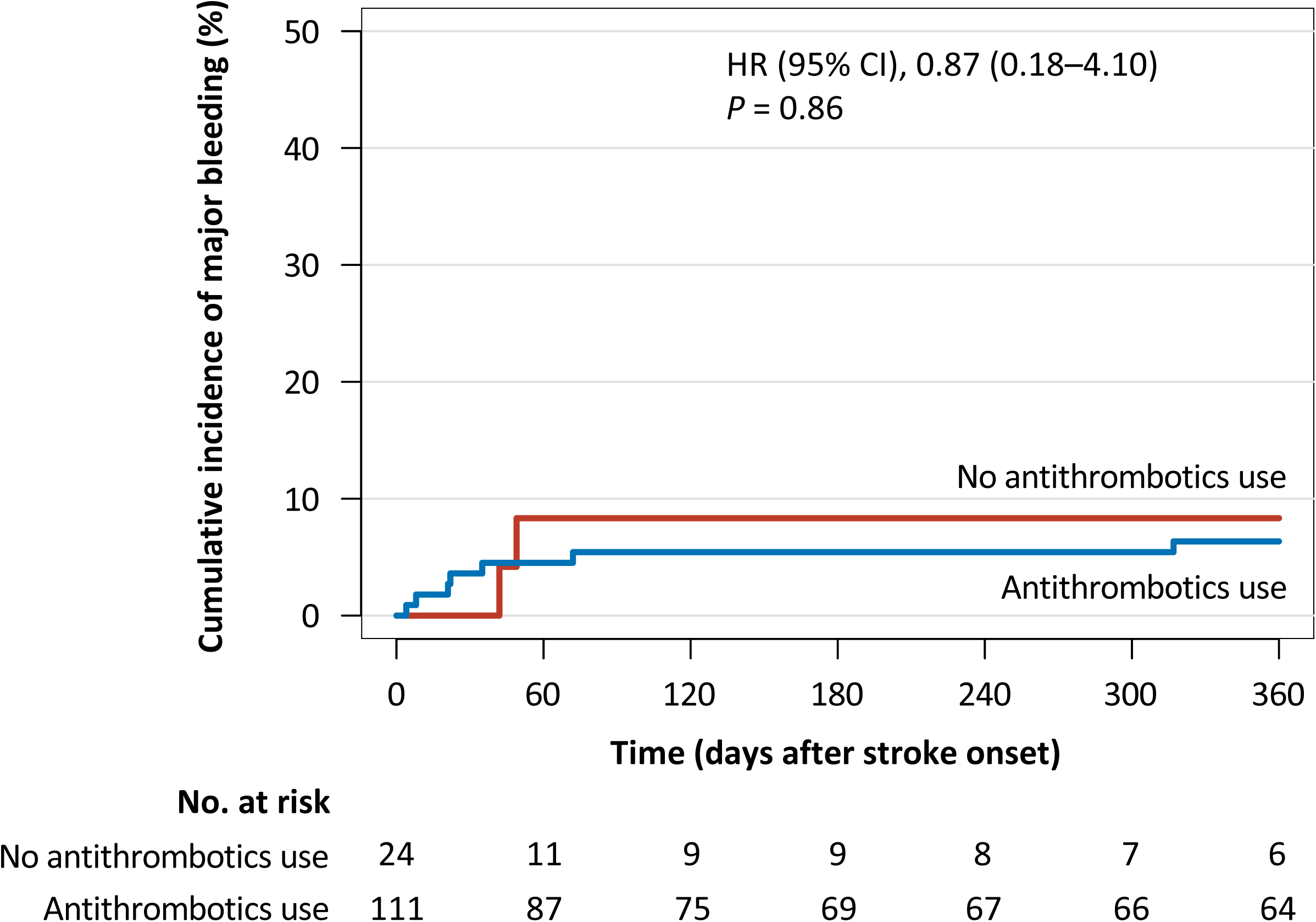
Cumulative incidence of major bleeding by antithrombotic agent use HR, hazard ratio; CI, confidence interval

### Survival Estimate Using Distant Metastatic Status, Stroke Subtype, and Plasma D-Dimer ***Levels***

Our analysis revealed that stroke mechanism, coagulation abnormalities, and cancer progression contributed to predict the prognosis in patients with ischemic stroke and active cancer. To investigate the usefulness of plasma D-dimer levels, we examined the relationship between plasma D-dimer levels and survival outcomes, stratified by distant metastasis and stroke subtypes.

**Figure 4** shows the distribution of the plasma D-dimer levels according to distant metastasis and cryptogenic stroke status. Plasma D-dimer levels were significantly higher in patients with metastasis than those with non-metastasis (**Figure 4A**, 8.51 [2.20–20.72] μg/mL vs. 1.91 [0.79–6.71] μg/mL, *P* < 0.001). As already shown in **Table 2**, patients with cryptogenic stroke had significantly higher plasma D-dimer levels than those with known etiologies (**Figure 4B**, 10.35 [3.59–22.38] μg/mL vs. 1.76 [0.83–3.73] μg/mL, *P* < 0.001).

**Figure 4.**
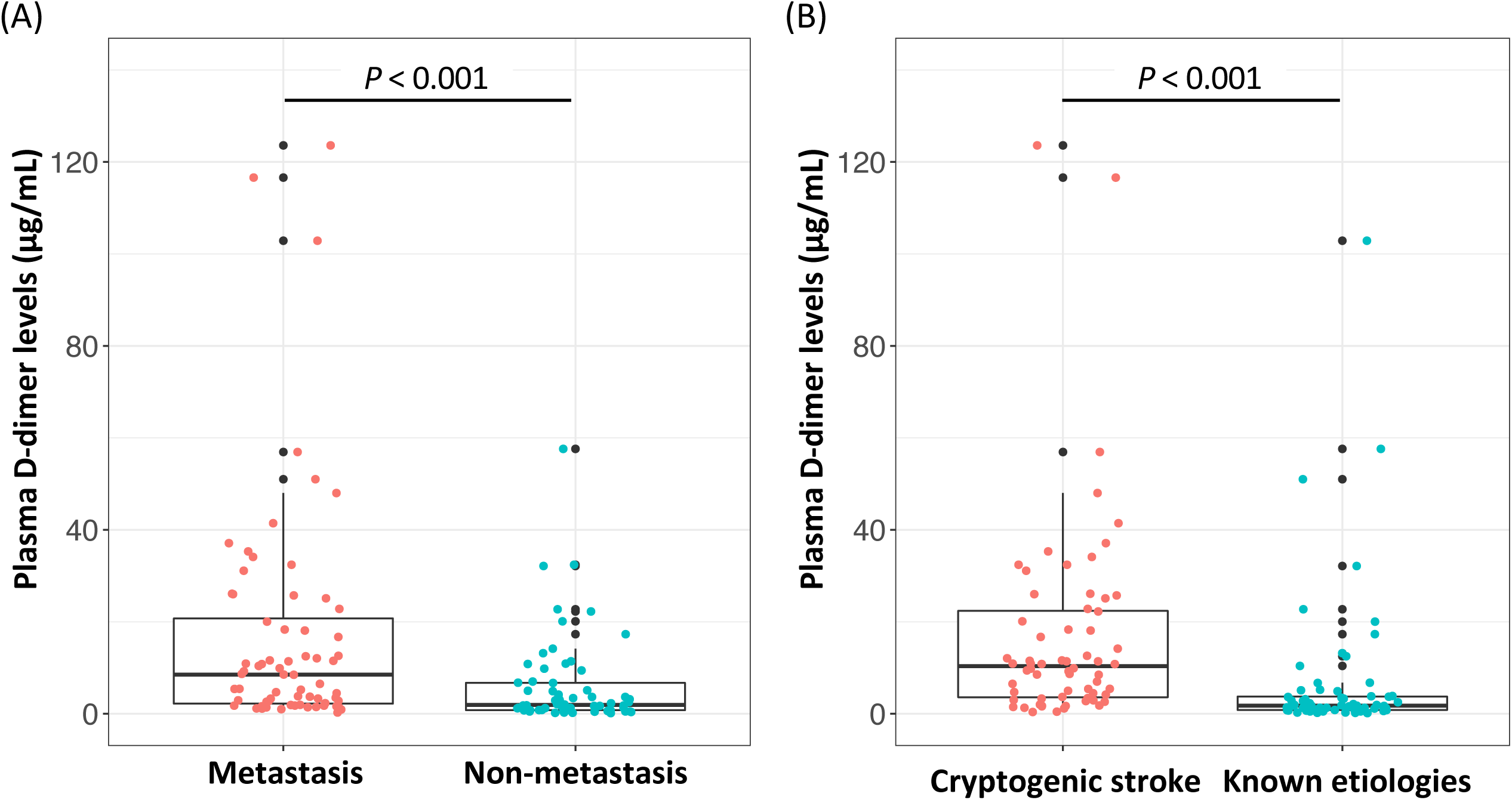
The distribution of plasma D-dimer levels by (A) distant metastasis and (B) stroke subtypes (A) The median plasma D-dimer levels for patients with metastasis was 8.51 μg/mL (interquartile range of 2.20–20.72 μg/mL), and that for patients with non-metastasis was 1.91 (interquartile range of 0.79–6.71 μg/mL). (B) The median plasma D-dimer levels for patients with cryptogenic stroke was 10.35 μg/mL (interquartile range of 3.59–22.38 μg/mL), and that for patients with known etiologies was 1.76 (interquartile range of 0.83–3.73 μg/mL).

**Figure 5A** shows the survival curves based on distant metastatic status and median plasma D-dimer levels of each group. As shown in **Figure 4A**, the median plasma D-dimer levels were 1.91 μg/mL for the group with non-metastasis and 8.51 μg/mL for the group with metastasis; therefore, we divided the patients into four groups based on distant metastasis status and median plasma D-dimer levels. The 1-year survival rates were 12% in patients with distant metastasis and high plasma D-dimer levels and 84% in patients with non-metastasis and low plasma D-dimer levels. **Figure 5B** shows the survival curves based on stroke subtype and median plasma D-dimer levels of each group. As shown in **Figure 4B**, the median plasma D-dimer levels were 1.76 μg/mL for the known etiologies group and 10.35 μg/mL for the cryptogenic stroke group; therefore, we divided the patients into four groups based on the stroke subtype and median plasma D-dimer levels. The 1-year survival rates were 20% in patients with cryptogenic stroke and high plasma D-dimer levels and 81% in patients with known stroke etiologies and low plasma D-dimer levels. These results suggest that plasma D-dimer levels on admission provide relevant information for predicting the prognosis of patients with ischemic stroke and active cancer.

**Figure 5.**
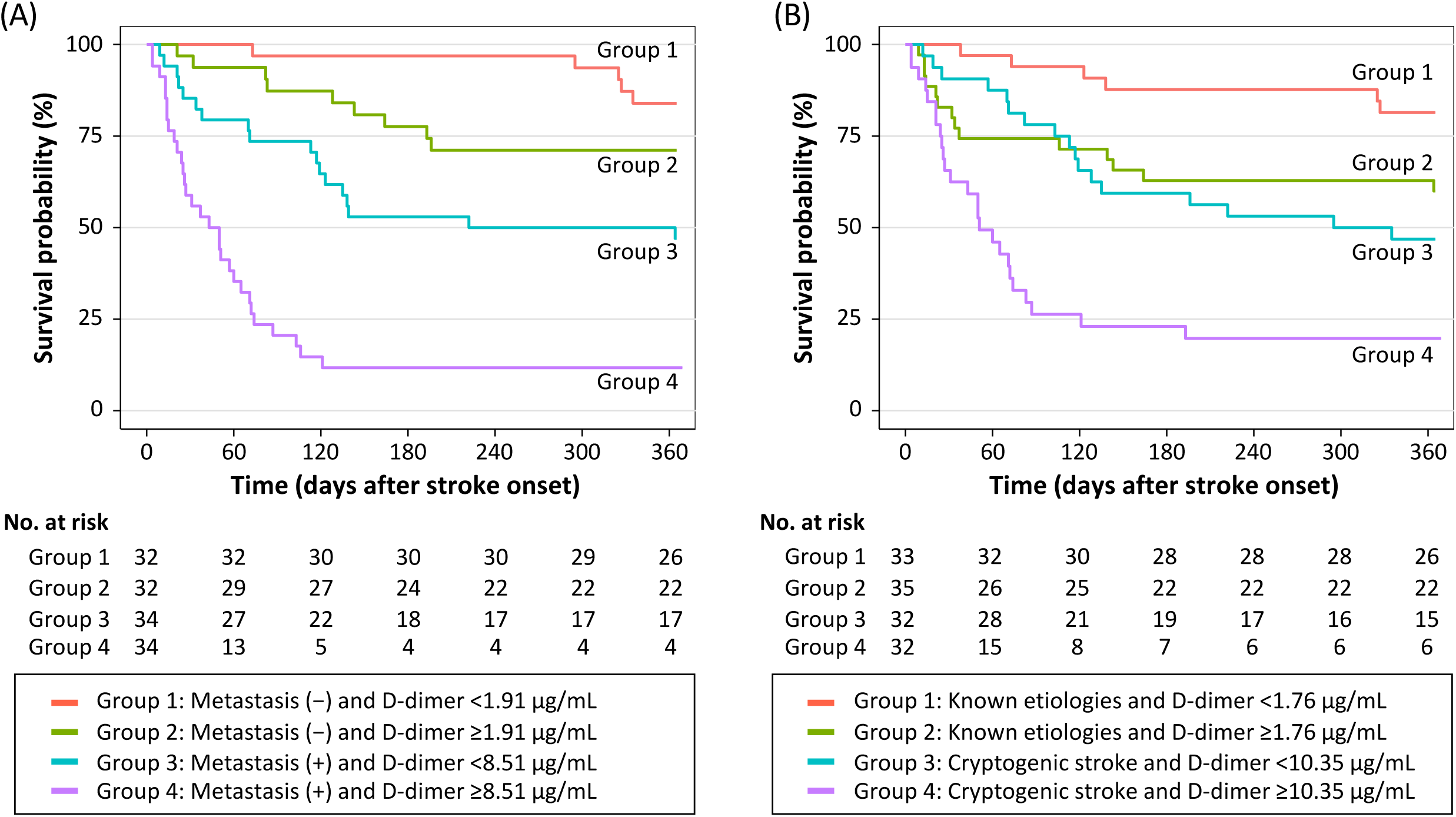
Kaplan-Meier survival curves stratified by (A) distant metastasis and (B) stroke subtype based on quartiles of plasma D-dimer levels We analyzed the survival rates of 132 patients since the plasma D-dimer levels on admission were not available for three patients. (A) The median plasma D-dimer levels were 1.91 μg/mL for the group with non-metastasis and 8.51 μg/mL for the group with metastasis; therefore, we divided the patients into four groups based on distant metastasis status and median plasma D-dimer levels. (B) The median plasma D-dimer levels were 1.76 μg/mL for the known etiologies group and 10.35 μg/mL for the cryptogenic stroke group; therefore, we divided the patients into four groups based on stroke subtype and median plasma D-dimer levels. Plasma D-dimer levels provide additional prognostic information for patients with distant metastasis and stroke subtype.

## Discussion

We investigated the prognosis of patients with ischemic stroke and active cancer using data from the SCAN study, a prospective, multicenter, observational study in Japan. Patients with cryptogenic stroke had shorter survival times than those with known etiologies. Distant metastasis, plasma D-dimer levels, and DVT/PE complications at stroke onset were independently associated with mortality after adjusting for potential confounders. Furthermore, plasma D-dimer levels stratified the prognosis of patients with distant metastasis and cryptogenic stroke. The current study suggests that there is a strong relationship between the degree of cancer-associated coagulopathy and the prognosis of patients with ischemic stroke and active cancer.

Previous studies have examined the prognosis of patients with ischemic stroke and active cancer. The median survival of these patients, considering all stroke subtypes, ranged from 84 to 109 days [6,7]. In our study, survival time was favorable compared with that in previous reports. The differences in survival among studies could owe to differences in the patients’ backgrounds. For example, the proportion of distant metastasis was 66% in the study by Lee et al., and 69% in the study by Navi et al., and 51% in our study. Additionally, cryptogenic stroke, a predictor of poor outcome in patients with ischemic stroke and active cancer, accounted for 72% in the study by Lee et al., and 51% in the study by Navi et al., and 48% in our study. Thus, differences in cancer progression and stroke subtypes may influence patients’ survival.

In our cohort, patients with cryptogenic stroke had shorter survival times than those with known etiologies. This result confirmed the finding from a previous retrospective study by Navi et al [8]. There are several possible explanations for this finding. First, patients with cryptogenic stroke and active cancer often have cancer-related coagulation abnormalities, resulting in poor outcomes [27]. This is supported by the finding that plasma D-dimer levels were significantly higher in patients with cryptogenic stroke than those with known etiologies [19–21]. Second, distant metastasis was more common in the cryptogenic stroke group, leading to poor prognosis. These backgrounds are associated with high mortality in patients with cryptogenic stroke and active cancer. The current study also demonstrated that patients with cryptogenic stroke had higher serum hsCRP levels than those with known etiologies. Cancer-related chronic inflammation could be associated with cryptogenic stroke and a poor prognosis [28].

Cryptogenic stroke has been reported as an independent predictor of mortality in patients with ischemic stroke and active cancer [8]. Other studies have shown plasma d-dimer levels to be also associated with poor prognosis [12,17,18]. Of note, plasma D-dimer levels are reportedly high in patients with cryptogenic stroke and active cancer [19–21]; hence, these findings may reflect underlying coagulation abnormalities. The present study examined the prognostic impact of cryptogenic stroke and plasma D-dimer levels using the multivariate model. Cryptogenic stroke was associated with poor prognosis in univariate analysis but was not significant in multivariate analysis. As mentioned earlier, plasma D-dimer levels were higher in patients with cryptogenic stroke than in those with known etiologies; therefore, the result that cryptogenic stroke was associated with death may be the effect of coagulation abnormalities. Interestingly, among patients with distant metastasis and/or cryptogenic stroke, those with high plasma D-dimer levels had poorer prognosis than those without. This finding supports the notion that the degree of cancer-associated coagulation abnormalities is related to prognosis. In clinical practice, it would be very helpful to use plasma D-dimer levels, in addition to distant metastasis and cryptogenic stoke, to predict the prognosis.

Regarding stroke subtypes, this study classified cryptogenic strokes as patients whose cause could not be confidently identified or had no apparent cause other than malignancy. However, the etiologic classification for patients with cancer-associated hypercoagulability is debatable. This is due to the diversity of cancer-associated stroke phenotypes. For example, if marantic endocarditis, also known as non-bacterial endocarditis, is detected by echocardiography in the cancer population, it would be classified as cardioembolism [24]. If blood tests confirm disseminated intravascular coagulation, it will be categorized as another determined etiology [24]. There are also cases of tumors or air embolisms [29,30]. Meanwhile, even if these causes were not found by screening, they may have gone undetected. If blood tests are incomplete, disseminated intravascular coagulation may not be diagnosed. Therefore, consensus about the etiologic classification for patients with cancer-associated hypercoagulability will be needed.

Patients with known etiologies and low D-dimer levels had a better prognosis than those with cryptogenic stroke and severe coagulation abnormalities. This means those with clear traditional stroke etiologies are likely cancer-independent strokes and have a much better prognosis than those with cancer-related strokes. This is presumably because cancer-associated coagulation abnormalities do not affect morbidity and mortality. In other words, patients with active cancer just happened to develop an ischemic stroke, and it is unlikely that cancer-associated hypercoagulability is related to the development of strokes. These results can be a message that clinicians should not deny stroke therapy based on a cancer history, where there is no reason to think cancer caused the stroke.

Hemorrhagic complications are among the most significant concerns in the antithrombotic treatment of ischemic stroke patients with active cancer [31]. In our analysis, antithrombotic therapy significantly reduced recurrent stroke and did not increase major bleeding. Because the SCAN study was observational, the indication for antithrombotic medications was left to the attending physician’s judgment. Therefore, it is likely that antithrombotic treatment was not given to patients at high risk of bleeding. However, our data prove that antithrombotic medications should be given to patients whose physicians determine they can use antithrombotic drugs. Further studies are needed to determine the pros and cons of antithrombotic therapy in stroke patients with active cancer.

The strength of this study resides in its prospective and multicenter design. However, this study had several limitations. First, although we tried to enroll as many patients as possible, we could not obtain consent from all the patients to participate in the study. It is possible that consent was not obtained from patients with advanced cancer and a selection bias could exist. Second, this study did not evaluate whether lowering plasma D-dimer levels with antithrombotic therapy improves prognosis. A recent study has reported that decreasing plasma D-dimer levels with antithrombotic use are associated with prognosis [18]. Third, events, such as stroke recurrence and major bleeding, were collected based on the history provided by the patient or family. Thus, the events may have been underestimated if they were not reported by the participants. Lastly, this study was limited to Japanese patients, so our findings may not generalize to other stroke populations.

In conclusion, this study analyzed the survival of patients with ischemic stroke and active cancer, using data from the SCAN study. The prognosis of ischemic stroke patients with active cancer varies considerably depending on distant metastasis, venous thromboembolism complications, and coagulation abnormalities. Based on this information, clinicians can decide how to treat patients with ischemic stroke and active cancer.

## Data Availability

Data are available on reasonable request to the corresponding author.

## Non-standard Abbreviations and Acronyms

HR: Hazard ratio
CI: Confidence interval
SCAN: Ischemic Stroke in Patients with Cancer and Neoplasia
NIHSS: National Institute of Health Stroke Scale
DVT: Deep venous thrombosis
PE: Pulmonary embolism
mRS: modified Rankin Scale
rt-PA: Recombinant-tissue plasminogen activator
hsCRP: High-sensitivity C-reactive protein
TOAST: Trial of Org 10172 in Acute Stroke Treatment
IQR: Interquartile range
AIC: Akaike’s information criterion

## Acknowledgements

We would like to thank all investigators for their efforts in conducting the SCAN study.

